# Circulating microRNA profiles in early-stage osteoarthritis and rheumatoid arthritis

**DOI:** 10.1101/2024.12.10.24318813

**Authors:** Madhu Baghel, Thomas Wilson, Michelle Ormseth, Patrick Yousif, Ayad Alkhatib, Alireza Meysami, Jason Davis, Vasilios Moutzouros, Shabana Amanda Ali

## Abstract

**Background:** Osteoarthritis (OA) and rheumatoid arthritis (RA) are prevalent forms of arthritis. Early detection of OA and RA is challenging with existing methods, which can delay effective management. MicroRNAs are small molecules that have emerged as promising disease biomarkers with the potential to improve early detection and differentiation of arthritis subtypes. In this study we aimed to identify distinct circulating microRNAs in plasma from individuals with early OA and early RA, using an unbiased microRNA-sequencing approach.

**Methods:** Plasma samples were collected from three study groups including: (a) early OA (N=20), individuals with knee OA symptoms and radiographic Kellgren-Lawrence grade 0 or 1; (b) early RA (N=12), treatment-naïve individuals with <6 months of RA symptoms in any joint; and (c) non-OA/RA (N=44), individuals with no history of arthritis. Of these, N=62 samples were subjected to microRNA-sequencing and analysis using a previously optimized pipeline. Exploratory analyses were followed by a stepwise filtering approach to shortlist both known (documented in miRBase v22.1) and novel (predicted using bioinformatics) microRNAs. Prioritized microRNAs were then validated via real-time qPCR (RT-qPCR) in N=14 independent samples.

**Results:** Principal component analyses revealed clustering of early OA versus both early RA and non-OA/RA groups, but not between early RA and non-OA/RA. In early OA, n=170 differentially expressed (DE) microRNAs were identified compared to both early RA and non-OA/RA, while no significant differences were found between early RA and non-OA/RA. Of these DE microRNAs, stepwise filtering and RT-qPCR validation identified dysregulation of six known microRNAs between early OA and early RA. Of these six microRNAs, two were upregulated in early OA, including hsa-miR-16-5p and hsa-miR-29c-3p, and four were upregulated in early RA, including hsa-miR-744-5p, hsa-miR-382-5p, hsa-miR-3074-5p, and hsa-miR-11400. Additionally, one novel microRNA sequence was found to be enriched in early OA and four in early RA.

**Conclusion:** We identified a total of six known and five novel circulating microRNAs that differ between early OA and early RA individuals. Validation of these microRNAs in independent cohorts is warranted to establish their biomarker potential for distinguishing individuals with early OA versus early RA.

## Background

Arthritis is a leading cause of joint pain and disability that encompasses over 100 subtypes (1). Among these, osteoarthritis (OA) and rheumatoid arthritis (RA) are the most common, with a global prevalence of 6.5% and 0.5-1%, respectively (2–4). Both conditions can affect a range of joints, from small phalangeal joints to larger weight-bearing joints, and both are driven by a multifactorial etiology involving genetic, epigenetic, and environmental factors (5, 6). OA is the most common type of non-autoimmune arthritis, primarily characterized by tissue destruction (5, 7). RA is the most common type of autoimmune arthritis, primarily characterized by joint inflammation (8). Although the pathophysiology of OA and RA differs, the initial presentation (e.g., symptoms such as joint pain) can overlap significantly, particularly in cases of single-joint disease (9–11). This creates a challenge for accurate diagnosis which is important in early stages of disease to enable timely implementation of appropriate interventions, including exercise for OA (12) and disease-modifying anti-rheumatic drugs (DMARDs) for RA (13).

Currently, diagnosis and staging criteria for OA rely on imaging, such as the Kellgren-Lawrence (KL) radiographic grading scale (11). For RA, diagnosis involves four domains as outlined by the American College of Rheumatology: serological testing [e.g., for rheumatoid factor (RF) and anti-citrullinated protein antibody (ACPA)], physical examination for joint swelling and tenderness, evaluation of symptom duration, and testing acute phase reactants like C-reactive protein (CRP) (14, 15). While these diagnostic methods are effective for established OA and RA, respectively, they are less reliable for detecting early-stage disease. Previous studies have defined early OA as individuals with KL grades 0 or 1 plus frequent joint pain lasting at least 4 weeks (16, 17), while early RA has been defined as treatment-naïve (i.e., no DMARD use) individuals with symptoms for <6 months and ACPA of an average 231.1 U/mL (range 24-2613.5 U/mL) (18). However, radiographic diagnosis of OA is dependent on identifying structural changes that are less apparent in the early stages (11). For RA, serological markers like CRP lack specificity for RA-related inflammation and can be elevated in other conditions such as systemic lupus erythematosus or psoriatic arthritis (19, 20). Given these challenges, there is a need for biomarkers that can reliably detect and distinguish OA and RA in the early stages.

MicroRNAs have emerged as promising candidate biomarkers (21). These small non-coding RNA molecules are abundant and stable in peripheral blood, exhibit resistance to degradation, and show disease- and stage-specific profiles (22). Although previous studies have identified microRNAs involved in both OA and RA (23–25), to date only three studies have explored circulating microRNAs in early OA or early RA individuals. Briefly, Ali et al. compared early to late OA plasma utilizing microRNA-sequencing (16), Romo-García et al. compared early RA to healthy control serum with microRNA microarray (18), and Cunningham et al. compared “at risk” RA to healthy control serum using a microRNA multiplex panel (26). To the best of our knowledge, no studies have compared circulating microRNAs in early OA and early RA alongside non-OA/RA.

Advances in sequencing technology and bioinformatic tools have made microRNA-sequencing the current gold standard approach for unbiased and comprehensive microRNA profiling (27). Compared to methods such as microarray, sequencing enables high-throughput identification of microRNAs and discovery of novel microRNA transcripts (28, 29). Novel microRNAs are detected based on their predicted secondary structures and defined based on specific criteria (e.g., a lack of sequence homology with other species) (30). As a result, novel microRNAs can be highly specific to the biological contexts in which they are identified (29, 31). In this study, we aimed to use sequencing and bioinformatics to identify known and novel circulating microRNA signatures in early OA and early RA individuals.

## Methods

### Study group characteristics

An overview of the study groups is shown in **Figure 1A**. Plasma samples were obtained from the Henry Ford Health (HFH) OA Biobank, Nashville VA Medical Center, and the Osteoarthritis Initiative (OAI, https://nda.nih.gov/oai) for both microRNA-sequencing and validation experiments. We used established definitions for early OA (16, 17) and early RA (18). The early OA group included individuals with symptoms and knee KL grades of 0 or 1, as assessed by an orthopedic surgeon. Of N=20 early OA samples, N=12 were used for sequencing and N=8 for validation. The early RA group included treatment-naïve individuals with <6 months of RA symptoms in any joint, as assessed by a rheumatologist. The measured ACPA levels in this group were 42 to 1196 U/mL, which aligns with the previously reported range of 24 to 2613.5 U/mL for early RA (18). Of N=12 early RA samples, N=6 were used for sequencing and N=6 for validation. The non-OA/RA group was obtained from the OAI “Control Cohort” and included individuals with no features of OA or RA. All N=44 non-OA/RA samples were used for sequencing. Clinicodemographic variables were collected as shown in **Table 1**, including age, sex, race, and body mass index (BMI). Institutional Review Board (IRB) approval was obtained from HFH (IRB #13995), the Nashville VA Medical Center (IRB #1643056 and IRB #1722175), and the OAI via the University of California, San Francisco (FWA approval # 00000068; IRB approval # 10-00532). Written informed consent was obtained from all participants before enrollment.

**Figure 1.**
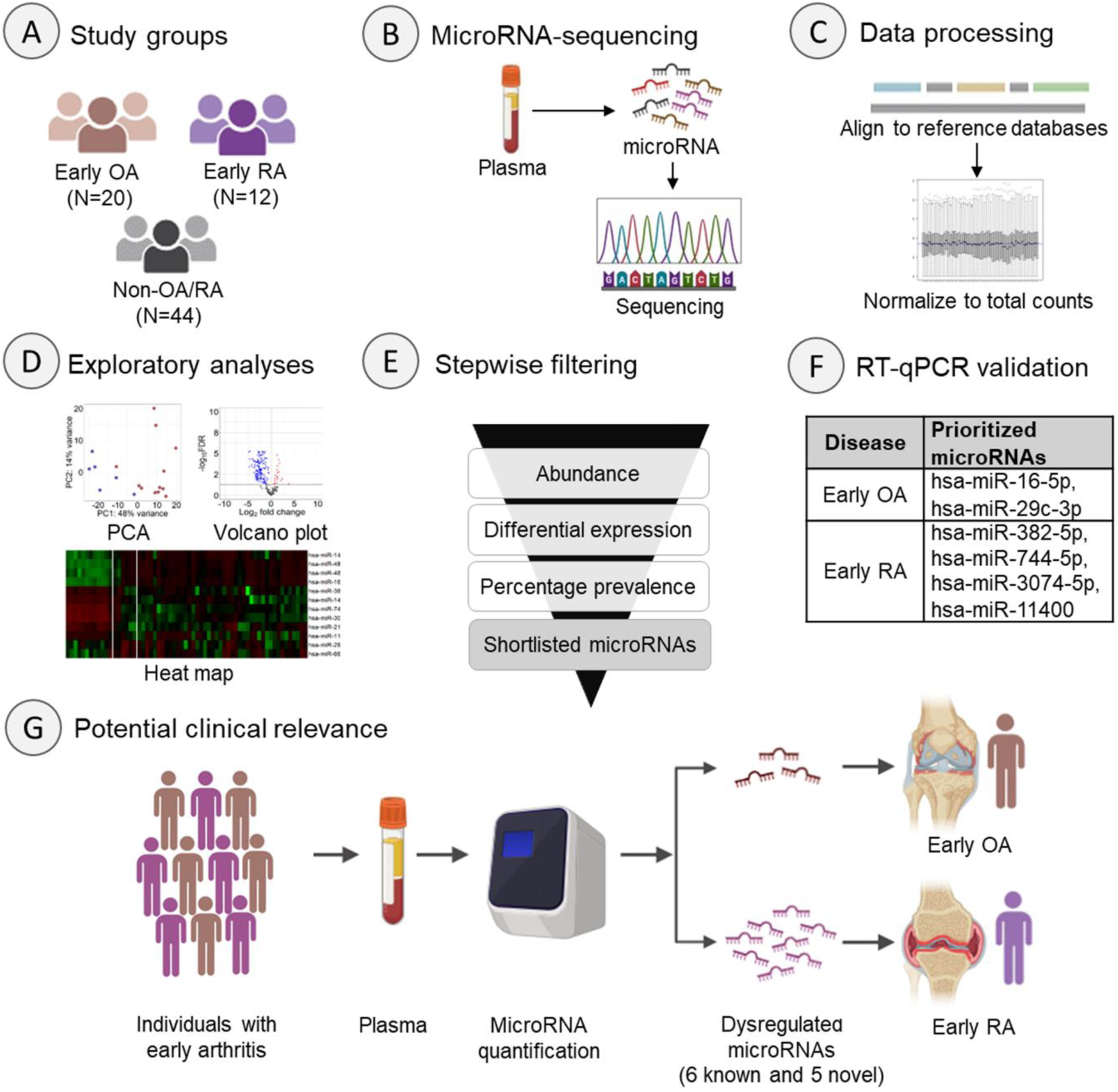
Overview of study design. (A) The study included three groups defined by predetermined inclusion and exclusion criteria. (B) MicroRNA-sequencing was performed on N=62 plasma samples. (C) Data were processed using an established pipeline (31). (D) Exploratory analyses included principal component analysis (PCA). (E) Stepwise filtering was performed to shortlist microRNAs. (F) Validation experiments in N=14 independent samples prioritized six microRNAs. (G) Dysregulated known and novel microRNAs may be used to distinguish individuals with early OA or early RA.

**Table 1.**
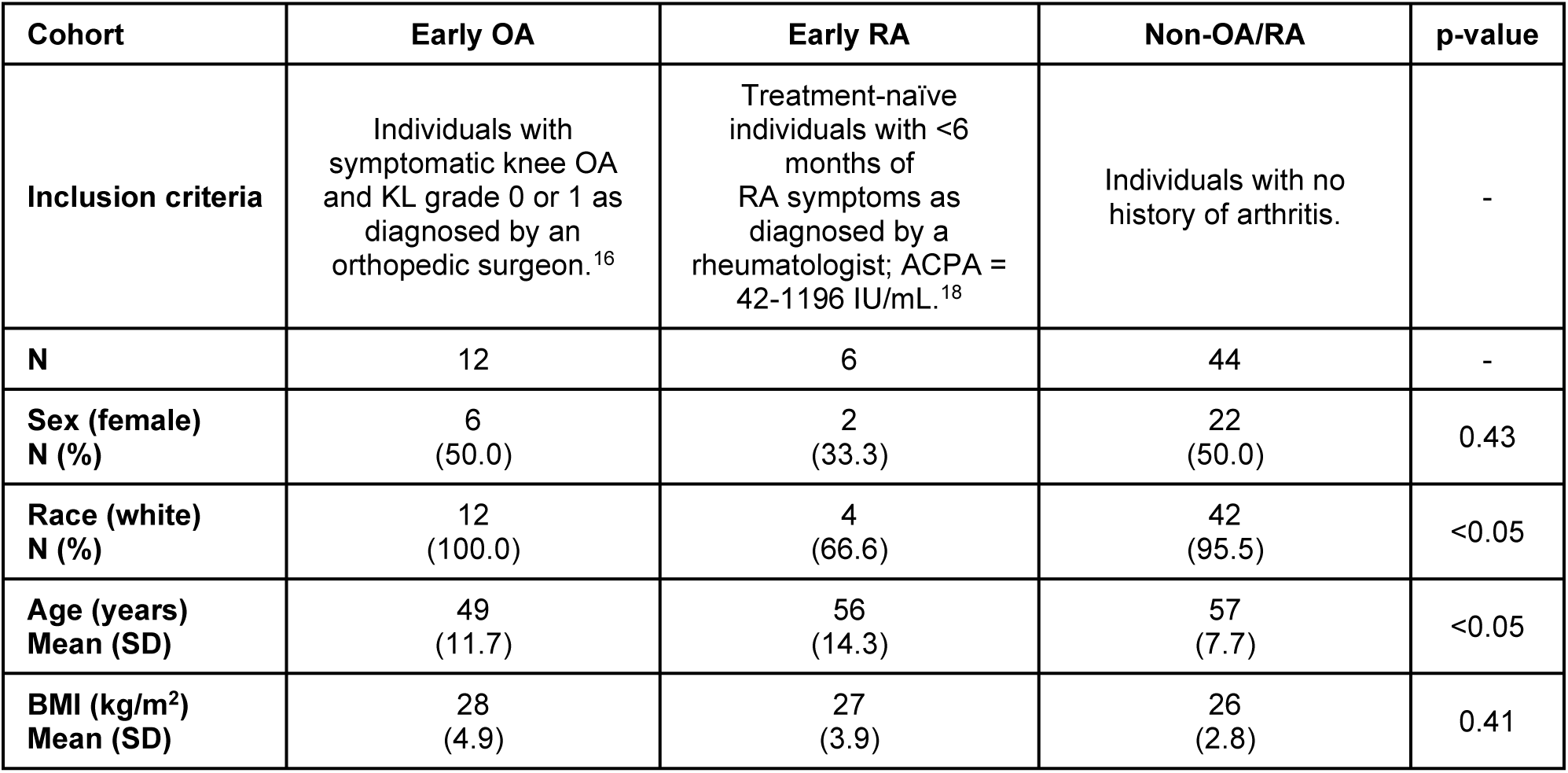
Characterization of the three study groups used for microRNA-sequencing. P-value shows the significance between groups assessed by one-way ANOVA for continuous variables and Chi-square test for categorical variables. KL = Kellgren-Lawrence; ACPA = Anti-citrullinated protein antibody; SD = standard deviation; BMI = body mass index.

| Cohort | Early OA | Early RA | Non-OA/RA | p-value |
| --- | --- | --- | --- | --- |
| **Inclusion criteria** | Individuals with symptomatic knee OA and KL grade 0 or 1 as diagnosed by an orthopedic surgeon.<sup>16</sup> | Treatment-naïve individuals with <6 months of RA symptoms as diagnosed by a rheumatologist; ACPA = 42-1196 IU/mL.<sup>18</sup> | Individuals with no history of arthritis. | - |
| **N** | 12 | 6 | 44 | - |
| **Sex (female)**<br>**N (%)** | 6<br>(50.0) | 2<br>(33.3) | 22<br>(50.0) | 0.43 |
| **Race (white)**<br>**N (%)** | 12<br>(100.0) | 4<br>(66.6) | 42<br>(95.5) | <0.05 |
| **Age (years)**<br>**Mean (SD)** | 49<br>(11.7) | 56<br>(14.3) | 57<br>(7.7) | <0.05 |
| **BMI (kg/m<sup>2</sup>)**<br>**Mean (SD)** | 28<br>(4.9) | 27<br>(3.9) | 26<br>(2.8) | 0.41 |

### Plasma collection

Blood samples were collected in EDTA-coated vacutainers and fractionated by centrifugation as previously described (16). Plasma was aliquoted into 250 μL per cryovial, flash frozen with liquid nitrogen, and stored at -80°C until use. RNA was extracted from plasma using the miRNeasy Serum/Plasma Advanced Kit (QIAGEN Inc.) following the manufacturer’s guidelines. ACPA was quantified in diluted plasma (dilution factor = 4), using the Human ACPA ELISA kit (abx257273; Abbexa LLC), following the manufacturer’s guidelines. Absorbance was measured at 450 nm using a plate reader (PerkinElmer).

### MicroRNA-sequencing

MicroRNA-sequencing and data analysis were performed for N=62 samples, according to previously published protocols (16, 28, 31). Briefly, RNA isolated from plasma was sequenced using a 75-base single-end read protocol at a depth of approximately 17 ± 2.5 million reads/sample on an Illumina NextSeq2000 sequencer (**Figure 1B**). Raw microRNA sequences were aligned to miRBase v22.1 (32) and the human reference genome (vGRCh38; **Figure 1C**). Next, we filtered for abundant microRNAs with at least 10 counts-per-million (CPM) in two or more samples, and normalized counts to total counts (16, 28). Differential expression analysis between groups was performed in a pairwise manner using a quasi-likelihood negative binomial model with trended dispersions with the edgeR package (v3.40.2; **Figure 1D,E**).

### Bioinformatics

Novel microRNA sequences were identified using miRDeep2 (30) as previously described (16, 31). Briefly, raw reads were aligned to the human reference genome (vGRCh38) with sequences included in the miRBase v22.1 human and hairpin microRNA databases excluded. Candidate novel microRNAs were subsequently filtered by significant randfold value (p<0.05), a measure of microRNA secondary structure stability, and by lack of homology with known mature microRNA sequences in mice. Prioritized novel microRNA sequences were subjected to NCBI nucleotide Basic Local Alignment Search Tool (BLASTN; https://blast.ncbi.nlm.nih.gov/Blast.cgi) against *Homo sapiens* sequences. The search considered 100% query cover, 100% identity match, and no set E-value.

### MicroRNA real-time polymerase chain reaction (RT-qPCR)

MicroRNA RT-qPCR was performed in N=8 early OA and N=6 early RA independent plasma samples (**Figure 1F**), as previously described (33, 34). Briefly, a poly(A) tail was added to the microRNA with poly(A) polymerase (M0276L; NEB), followed by reverse transcription (RT) for the synthesis of cDNA using poly(T)-containing RT primers. The resulting cDNA was amplified using a QuantStudio 7 Pro Real-Time PCR System (Applied Biosystems), with custom microRNA oligonucleotides (Sigma-Aldrich) as specified in **Supplemental Table 1** and SYBR Green dye (Bio-Rad). Hsa-miR-24-3p was used as an endogenous control as previously done (16, 28).

### Statistical analyses

All data analysis was performed using R statistical software (v.4.2.0). For microRNA discovery and validation, a power calculation estimated a minimum sample size of N=6/group to detect a differential expression threshold of fold-change ± 2, at 80% power. Principal component analysis (PCA) was performed on variance-stabilized transformed counts. Heatmaps were plotted using z-score-transformed CPM. RT-qPCR data were analyzed using the delta-delta-Ct method (35). Statistical significance was determined by false discovery rate (FDR) correction ≤0.05 for microRNA-sequencing, and two-tailed Student’s T-test for RT-qPCR analyses at p≤0.05.

## Results

### Exploratory analyses of microRNAs in early OA and early RA plasma

To discover circulating microRNAs dysregulated in either early OA or early RA (**Figure 1G**), we performed microRNA-sequencing on N=62 plasma samples as shown in **Table 1**. Samples were matched for clinicodemographic variables as closely as possible, though significant differences in age and race across groups were noted (**Table 1**). After sequencing, read alignment to miRBase and the human reference genome resulted in raw counts for 2656 mature microRNAs. Data analyses were performed in a pairwise manner, leading to three comparisons: (1) early OA versus early RA, (2) early OA versus non-OA/RA and (3) early RA versus non-OA/RA. First, we filtered for microRNAs with >10 CPM in two or more samples (within each comparison), resulting in 285 abundant microRNAs for early OA versus early RA, 364 for early OA versus non-OA/RA, and 354 for early RA versus non-OA/RA. Next, we performed PCA to identify components that account for the variance within the counts. PCA plots showed distinct clustering between early OA and both early RA (**Figure 2A**) and non-OA/RA (**Figure 2B**), while no clustering was observed between early RA and non-OA/RA (**Figure 2C**). Despite the significant differences in age and race across groups (**Table 1**), no clustering was observed when classifying samples by age, sex, BMI, or race (**Supplemental Figure 1**), suggesting that these variables did not significantly impact microRNA counts. Following this, we performed unsupervised hierarchical clustering of microRNA counts for each pairwise comparison. Using heatmaps to visualize the results, we observed patterns of high and low levels of microRNAs in both the early OA versus early RA (**Supplemental Figure 2A**) and early OA versus non-OA/RA comparisons (**Supplemental Figure 2B**); however, no appreciable patterns were observed in the early RA versus non-OA/RA comparison (**Supplemental Figure 2C**). Finally, we performed pairwise differential expression analyses to identify significant differences in microRNAs in each comparison. At FDR≤0.05, we identified 170 differentially expressed (DE) microRNAs between early OA and early RA (**Figure 2D**) and 305 DE microRNAs for early OA versus non-OA/RA (**Figure 2E**); however, no significant differences were found between early RA and non-OA/RA (**Figure 2F**). Taken together, these results suggest there are distinct circulating microRNAs in early OA versus both early RA and non-OA/RA, but not between early RA and non-OA/RA.

**Figure 2.**
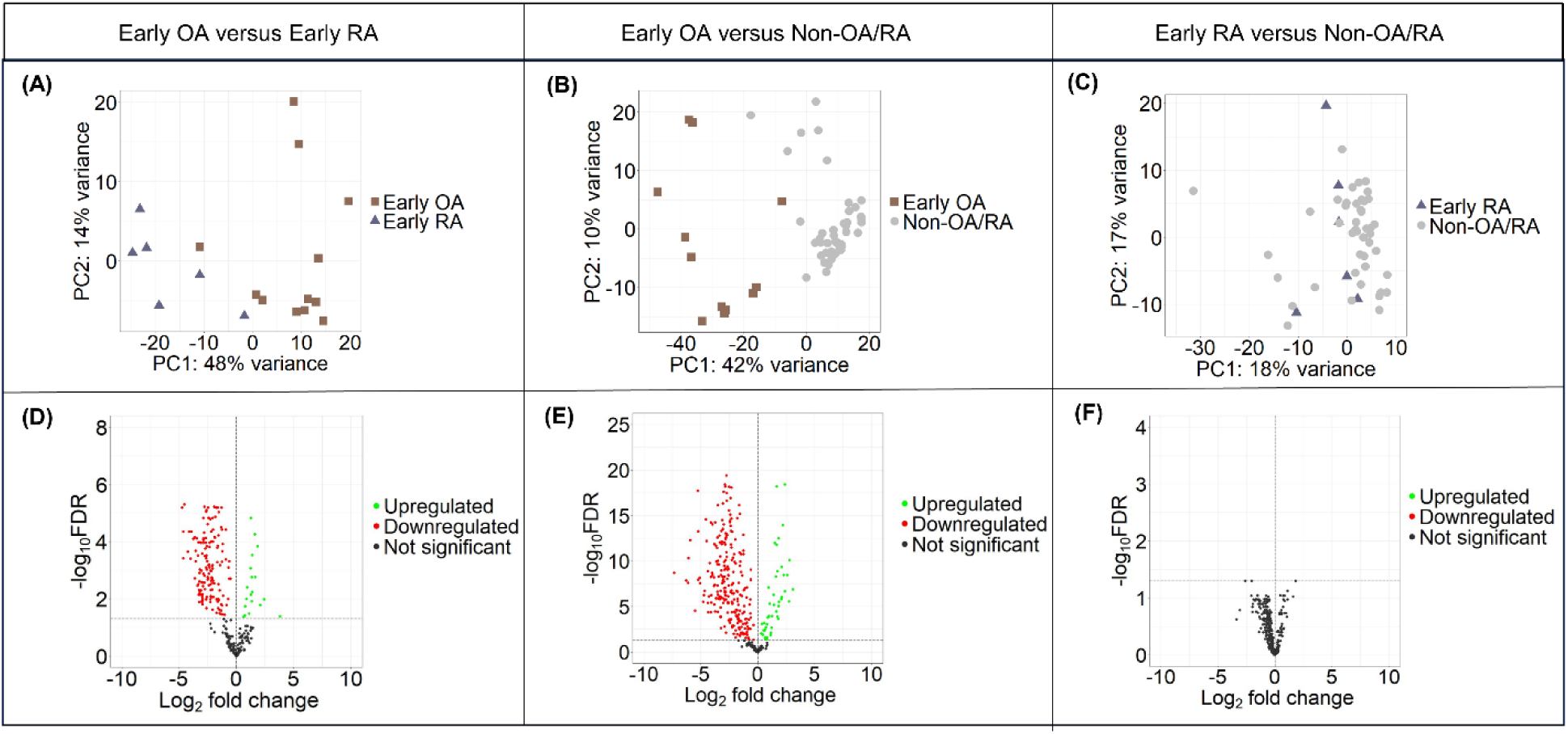
Distinct circulating microRNAs are present in early OA and early RA. (A-C) Principal component analyses showing clustering of samples when stratified by cohort. Each point represents a sample. Each axis represents the percentage variance for the principal component (PC). (D-F) Volcano plots showing differentially expressed microRNAs in each comparison, with FDR≤0.05 (horizontal line). Each dot represents a microRNA.

### Stepwise filtering of microRNAs dysregulated in early OA and early RA

To systematically identify dysregulated microRNAs associated with early OA and early RA, we implemented a stepwise filtering approach. The first steps retained abundant microRNAs (**Figure 3A, Criterion 1**) and DE microRNAs (**Figure 3A, Criterion 2**) for each pairwise comparison, as described above. We then compared the lists of DE microRNAs to identify those that were shared and found all 170 DE microRNAs identified in the early OA versus early RA comparison were also dysregulated in the early OA versus non-OA/RA comparison. Since no additional DE microRNAs were detected between early RA and non-OA/RA, we focused on the 170 DE microRNAs in early OA versus both early RA and non-OA/RA (**Figure 3A, Criterion 2**). Next, we filtered for microRNAs that were consistently up- or downregulated across samples in a group relative to their median levels in the comparison group (**Figure 3A, Criterion 3**). For early OA, 13 microRNAs were upregulated and 132 downregulated in 100% of samples compared to their median counts in early RA group. Of these, all 13 upregulated and 128 of the 132 downregulated microRNAs showed the same trend when compared to median counts in the non-OA/RA group (**Figure 3A, Criterion 3**). For early RA, 137 microRNAs were upregulated and 15 downregulated in 100% of samples versus their median counts in the early OA group. Since no microRNAs met the 100% threshold in the early RA versus non-OA/RA comparison, we relaxed this threshold to ≥83% of early RA samples and identified two upregulated (hsa-miR-11400, hsa-miR-21-5p) and two downregulated (hsa-miR-660-5p, hsa-miR-29c-3p) microRNAs (**Figure 3A, Criterion 3**). These four microRNAs represented our shortlisted panel for early RA (**Supplemental Table 2**). To refine the list of microRNAs for early OA, we performed an additional filtering step to select microRNAs with average logCPM ≥10 and logFC ≥ 1 or ≤ -2.5 in early OA versus early RA. This resulted in a shortlisted panel of four upregulated (hsa-miR-144-3p, hsa-miR-486-3p, hsa-miR-486-5p, hsa-miR-16-5p) and four downregulated (hsa-miR-744-5p, hsa-miR-382-5p, hsa-miR-3074-5p, hsa-miR-142-3p) microRNAs for early OA (**Figure 3A, Criterion 4; Supplemental Table 3**). In sum, this filtering approach shortlisted 12 microRNAs with distinct profiles in early OA and early RA plasma (**Figure 3B**).

**Figure 3.**
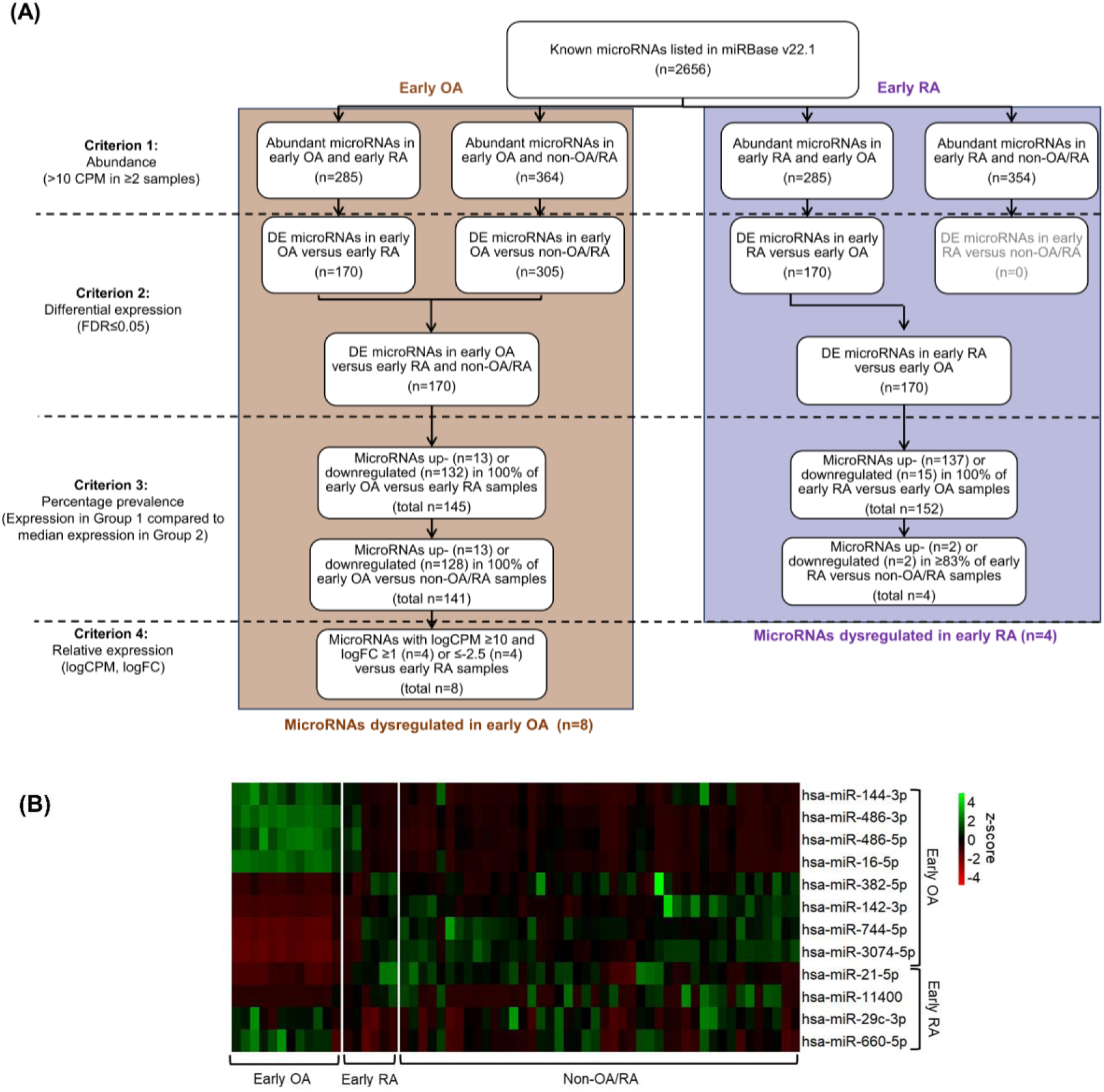
Stepwise filtering identified microRNAs dysregulated in early OA and early RA. (A) Four sequential filtering criteria were applied to identify dysregulated microRNAs. The brown box shows filtering for microRNAs in early OA versus early RA and non-OA/RA. The purple box shows filtering for microRNAs in early RA versus early OA and non-OA/RA. (B) Heatmap depicting plasma levels of 12 microRNAs dysregulated in early OA and early RA. Z-scores represent standardized microRNA counts, with green indicating higher levels and red indicating lower levels.

### MicroRNA validation in independent early OA and early RA samples

After shortlisting 12 microRNAs (**Figure 3**), we next sought to validate their levels in independent plasma samples using RT-qPCR. We focused on early OA versus early RA as our most clinically relevant comparison and measured the levels of each microRNA in N=8 early OA and N=6 early RA plasma samples (**Supplemental Table 4**). Of the 12 microRNAs, six showed patterns consistent with our microRNA-sequencing results. Among these, hsa-miR-16-5p and hsa-miR-29c-3p were elevated in early OA (**Figure 4A,B**), while hsa-miR-774-5p, hsa-miR-382-5p, hsa-miR-3074-5p, and hsa-miR-11400 were elevated in early RA (**Figure 4C-F**). The six remaining microRNAs either did not reach statistical significance or showed trends opposite to our sequencing data (**Supplemental Figure 3**). Overall, these results prioritize hsa-miR-16-5p, hsa-miR-29c-3p, hsa-miR-774-5p, hsa-miR-382-5p, hsa-miR-3074-5p, and hsa-miR-11400 as six circulating microRNAs that are different between early OA and early RA (**Figure 1F**).

**Figure 4.**
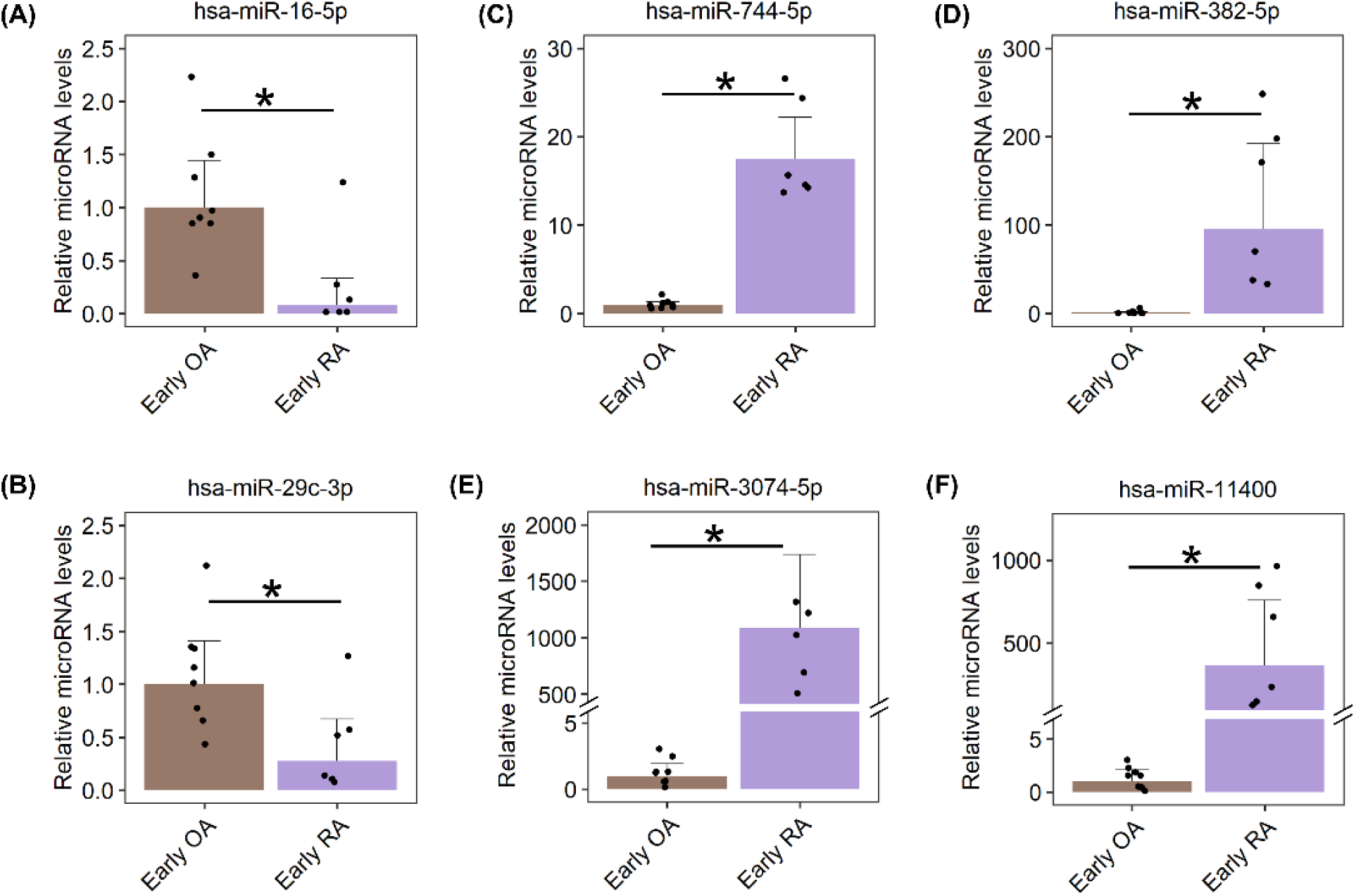
RT-qPCR validation of six microRNAs in early OA and early RA. RT-qPCR was performed in N=8 early OA and N=6 early RA independent plasma samples. Relative microRNA levels represent fold-change in early RA (purple) normalized to average levels in early OA (brown). *p<0.05; error bars = 95% confidence interval. (A,B) Two microRNAs were upregulated in early OA versus early RA. (C-F) Four microRNAs were upregulated in early RA versus early OA.

### Novel microRNA discovery in early OA and early RA

We next used established protocols to discover novel microRNAs in our sequencing dataset (16, 31). This approach identified 1257 unique predicted novel microRNA sequences that were present in at least one early OA or early RA sample, had a randfold p-value of <0.05, and showed no homology with known microRNA sequences in mice. To refine this list, we retained novel microRNAs present in ≥50% of either early OA or early RA samples, resulting in three and ten novel microRNAs, respectively (**Table 2**). BLASTN analysis confirmed the human origin of all 13 novel microRNA sequences, demonstrating 100% identity match and 100% query coverage when compared to *Homo sapiens* sequences. Looking for novel microRNAs that were enriched in early OA, we identified Novel miRNA EOA 2 in ≥75% of early OA samples and <25% of both early RA and non-OA/RA samples (**Table 2**). Meanwhile, four novel microRNAs (Novel miRNA ERA 2, 3, 4, and 7) were present in ≥50% of early RA samples, and completely absent in early OA samples (**Table 2**). We therefore prioritized these five novel microRNAs as the most distinct between early OA and early RA.

**Table 2.**
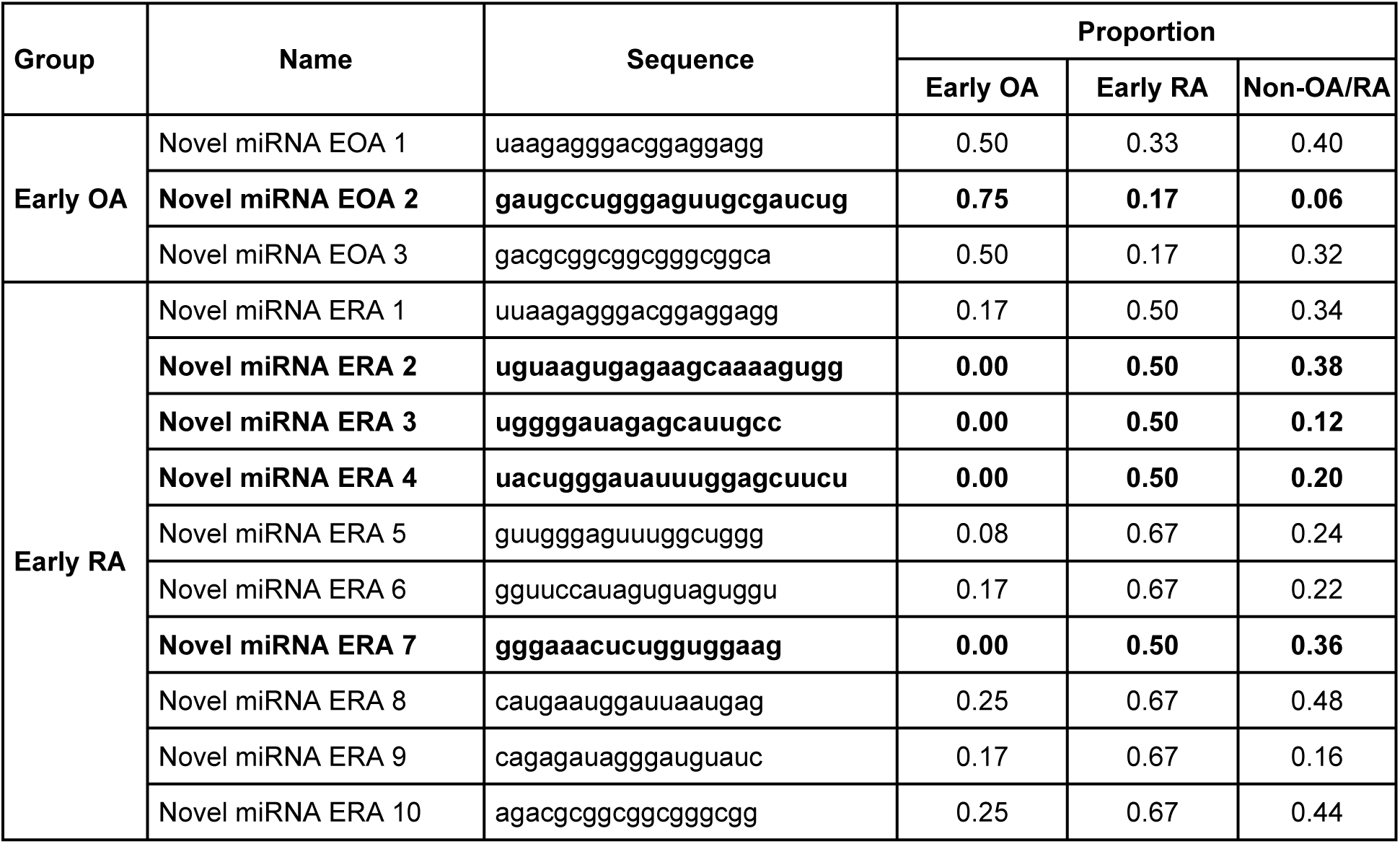
MicroRNA-sequencing identified five novel microRNAs enriched in early OA and early RA. List of all novel microRNA sequences found in ≥50% of early OA or early RA samples. Proportion = % of samples in each group that have the novel microRNA sequence. Bold = novel microRNAs prioritized for early OA and early RA.

## Discussion

Detecting early OA and early RA in clinical settings can be challenging since current diagnostic tools work best for established disease. Distinguishing early OA from early RA can also be challenging given the overlap in the initial presentation. These challenges point to the need for reliable, minimally invasive biomarkers to support accurate diagnosis and timely intervention. There is mounting evidence demonstrating the utility of non-coding RNAs as disease biomarkers (21, 25). In this study, we explore the potential of circulating microRNAs by performing microRNA-sequencing on plasma samples from individuals with early OA, early RA, and non-OA/RA. We observe distinct patterns of microRNAs between early OA and both early RA and non-OA/RA, but no significant differences between early RA and non-OA/RA. With stepwise filtering and real-time PCR validation, we prioritize a panel of six known microRNAs and five novel microRNAs as candidates to differentiate early OA from early RA.

Among the strengths of this study is the customized microRNA-sequencing analysis pipeline and systematic filtering approach we applied (16, 31). A common pitfall of profiling studies is the reliance on single thresholds, such as p-values or fold-changes, to select targets of interest. These thresholds can vary significantly between studies (36, 37), leading to irreproducible results; moreover, this strategy risks prioritizing biomarker candidates with statistical significance but not biological relevance. To overcome this, we designed a filtering approach to prioritize microRNAs with properties that would make strong biomarkers. First, we selected high abundance microRNAs to increase the likelihood their presence would be detectable in a general population. This is important because many microRNAs have low to no counts in a sequencing dataset, so while their fold change may be statistically significant, they may not play a biologically relevant role (38). Second, we performed pairwise differential expression analyses to consider each Group 1 to both Groups 2 and 3. This allowed us to prioritize microRNAs in early OA versus both early RA and non-OA/RA, and to find that microRNAs in early RA were different compared to early OA, not but non-OA/RA. Third, to counter the heterogeneity inherent to human biospecimens, we selected DE microRNAs that showed an increase or decrease across the majority of individual samples in a group. This allowed us to prioritize microRNAs with consistent trends and a greater likelihood of being reproducible. Interestingly, nearly all the microRNAs in the early OA group showed exclusive up- or downregulation compared to both early RA and non-OA/RA, suggesting a robust microRNA signature for early OA. As a fourth and final step to refine this early OA signature, we applied thresholds based on abundance and fold-change to retain microRNAs with the greatest difference between groups. Importantly, these filtering criteria were designed to prioritize microRNAs with biomarker potential, which means other criteria may be more appropriate for microRNA studies with different objectives.

Previous studies have reported circulating microRNAs in early OA and early RA, including seven plasma microRNAs associated with early OA (16) and seven serum microRNAs associated with early RA (18) or “at risk” RA (26). Notably, there was no overlap between these 14 microRNAs and the microRNAs shortlisted in the current study; however, 12 of the 14 microRNAs showed a significant difference in at least one of our three pairwise comparisons. From Ali et al. (16), seven microRNAs were elevated in early versus late radiographic knee OA plasma; among these, four were significantly downregulated in our early OA samples compared to both early RA and non-OA/RA (hsa-miR-191-3p, hsa-miR-199a-5p, hsa-miR-335-5p, and hsa-miR-671-3p). From Romo-García et al. (18), hsa-miR-361-5p was reported to be elevated in early RA serum compared to non-RA controls, and it was significantly upregulated in our early RA samples compared to early OA, but not to non-OA/RA. From Cunningham et al. (26), six microRNAs were upregulated in serum from “at risk” RA individuals (symptomatic with positive ACPA or RF but without clinical synovitis or raised CRP) versus healthy controls (26). Of these, five were found to be significantly upregulated in our early RA samples compared to early OA, but not to non-OA/RA. Overall, the inconsistencies between previous literature and our findings are not unexpected given the variations in study groups, sample types, and analytical approaches. This underscores the need for standardized methodologies in biomarker discovery.

Given their lack of previous documentation, novel microRNAs are inherently valuable as potential disease-specific biomarkers (30). Ali et al. (16) previously reported four novel microRNAs that were enriched in early OA. Of these, two sequences (novel_miRNA_2 and novel_miRNA_3) were found in our dataset but did not meet our filtering criteria, as they were present in <50% of early OA samples. To our knowledge, no studies to date have reported novel microRNAs in early RA. We present four novel microRNA candidates that merit further investigation as biomarkers for early RA, including characterization to verify their authenticity as microRNAs.

Relationships between circulating microRNAs and pathological processes have been reported in arthritis (39, 40), suggesting circulating microRNAs can reflect local disease status. In early OA plasma, we found elevated levels of hsa-miR-16-5p and hsa-miR-29c-3p. Previous studies have reported increased levels of hsa-miR-16 in OA plasma versus non-OA controls (41), and decreased levels in early RA serum versus established RA (42). It is also upregulated in serum from RA individuals following treatment with anti-inflammatory agents (43). In terms of tissue functions, hsa-miR-16-5p has mechanistic connections to cartilage degradation in OA (41). While hsa-miR-29c-3p has not been previously explored in OA or RA circulation, elevated levels are found in late OA versus early OA synovial fluid (44). These findings are not directly comparable to ours, but inverse trends between circulating and local microRNA levels have been previously reported (40, 45). In early RA plasma, we found elevated levels of hsa-miR-382-5p, hsa-miR-774-5p, hsa-miR-3074-5p, and hsa-miR-11400. Little is known about the role of these microRNAs in arthritis, with only one study examining the behavior of hsa-miR-382-5p in RA fibroblast-like synoviocytes (46). As such, the current study is the first to propose these four microRNAs as potential biomarkers for early RA.

To note the limitations of this study, our sample sizes were sufficient to detect significant microRNAs at FDR≤0.05 but were not sufficient to support the testing of predictive models (47, 48). Future studies with larger cohorts are required to evaluate the sensitivity and specificity of our prioritized microRNAs as biomarkers for early OA and early RA. Second, the microRNA signature we identify for early OA appears to be more robust than that identified for early RA, given the lack of differences between early RA and non-OA/RA. While previous studies report differences between early RA and healthy controls (18, 26), the utility of our study lies primarily in the comparison between early RA and early OA to ultimately support differentiation of these cases in clinical settings. Finally, we did not define OA by the number of joints involved, or RA by the specific joints involved, which may influence microRNA patterns. Future microRNA biomarker discovery studies are needed to compare single-joint knee OA versus RA, and RA versus other inflammatory conditions with overlapping serological markers.

## Conclusion

To our knowledge, this is the first study that profiles circulating microRNAs in early OA, early RA, and non-OA/RA individuals using microRNA-sequencing. With a tailored approach to data analysis, we prioritized 11 circulating microRNAs (six known and five novel) that are associated with early OA and early RA. These findings suggest there is potential for circulating microRNAs to serve as minimally invasive biomarkers for early OA and early RA, including as composite biomarkers on panels that include proteins. With further research in larger cohorts, these microRNAs can be developed into a diagnostic test to improve arthritis care.

## Data Availability

The microRNA-sequencing datasets generated during this study were deposited in the Gene Expression Omnibus database and will be released following peer-reviewed publication.

## List of abbreviations

ACPA: Anti-citrullinated protein antibody
BLASTN: nucleotide Basic Local Alignment Search Tool
BMI: Body mass index
CPM: Counts per million
CRP: C-reactive protein
DE: Differentially expressed
DMARDs: Disease-modifying anti-rheumatic drugs
FC: Fold-change
FDR: False discovery rate
KL: Kellgren-Lawrence
PCA: Principal component analysis
RA: Rheumatoid arthritis
RF: Rheumatoid factor
RT-qPCR: Real time polymerase chain reaction
OA: Osteoarthritis

## Declarations

### Ethics approval and consent to participate

All participants provided written informed consent to be included in this study. The study protocol followed guidelines set by the Henry Ford Health Institutional Review Board (IRB #13995). Additional ethics approval was provided by the VA Tennessee Healthcare System (IRB #1643056 and IRB #1722175), and the OAI via University of California, San Francisco (FWA approval # 00000068; IRB approval # 10-00532).

### Author contributions

MB, TGW, and SAA were involved in study conception and design. MB and TGW performed experimental work and data acquisition. JD, VM, MO, PY, AA, AM, MB and TGW contributed to biospecimen collection. MB, TGW, and SAA performed data analysis, interpretation, and drafted the manuscript. All authors contributed to manuscript revisions and approved the final version.

## Supplemental display items

**Supplemental Figure 1.**
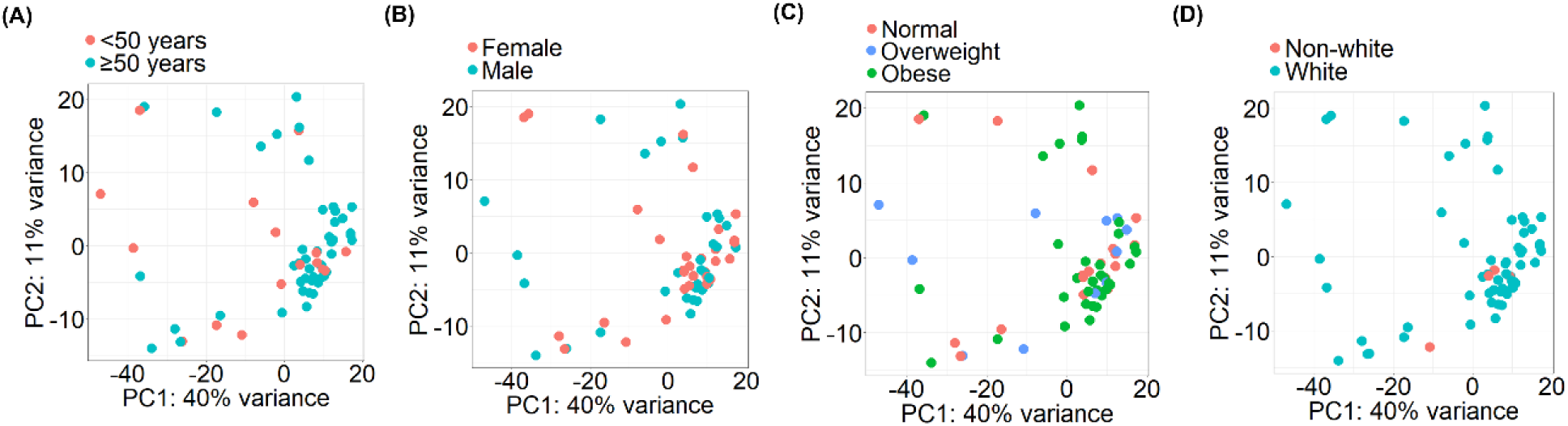
No distinct clustering is observed when microRNA-sequencing data are stratified by age, sex, BMI, or race. Principal component analysis plots of microRNA-sequencing data stratified by (A) age, (B) sex, (C) BMI (<25 kg/m^2^ = normal; 25-30 kg/m^2^ = overweight; >30 kg/m^2^ = obese), and (D) race. Each dot represents a sample. Each axis represents percentage variance for the principal component (PC).

**Supplemental Figure 2.**
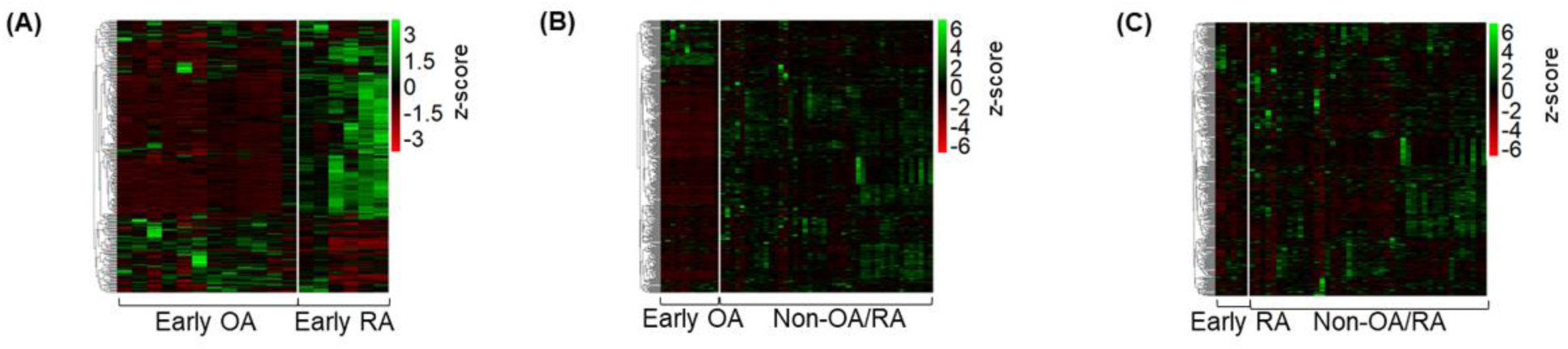
Expression profiles of microRNAs in pairwise comparisons. Heatmaps depicting unsupervised hierarchical clustering of microRNAs in individual samples of each pairwise comparison: (A) early OA versus early RA, (B) early OA versus non-OA/RA, and (C) early RA versus non-OA/RA. Z-scores represent standardized microRNA counts, with green indicating higher levels and red indicating lower levels.

**Supplemental Figure 3.**
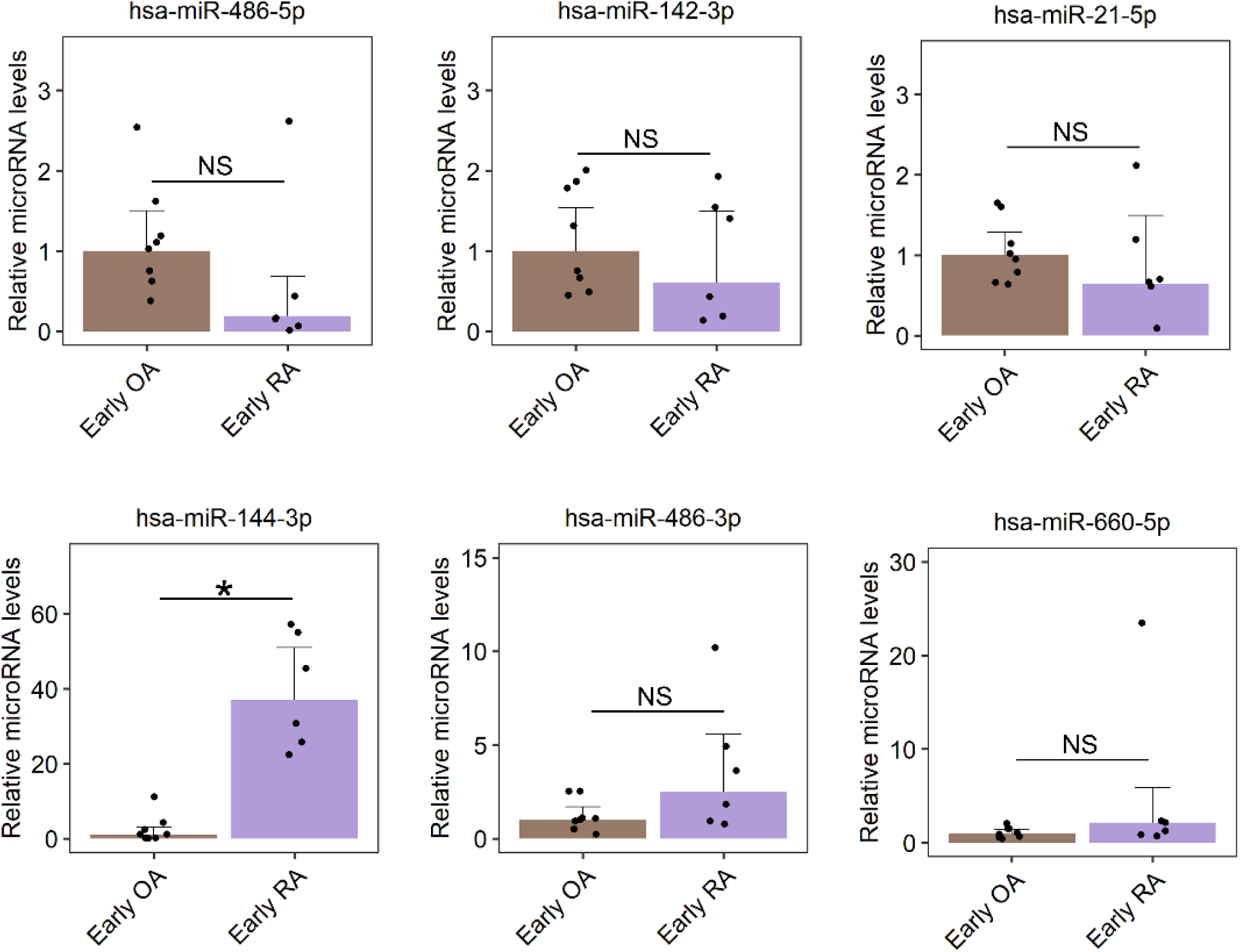
RT-qPCR for six microRNAs in early OA and early RA showing results inconsistent with sequencing data. RT-qPCR was performed in N=8 early OA and N=6 early RA independent plasma samples. Relative microRNA levels represent fold-change in early RA (purple) normalized to average levels in early OA (brown). Significance is denoted by a horizontal bar with *p<0.05 or NS = non-significant. Error bars = 95% confidence interval.

**Supplemental Table 1.**
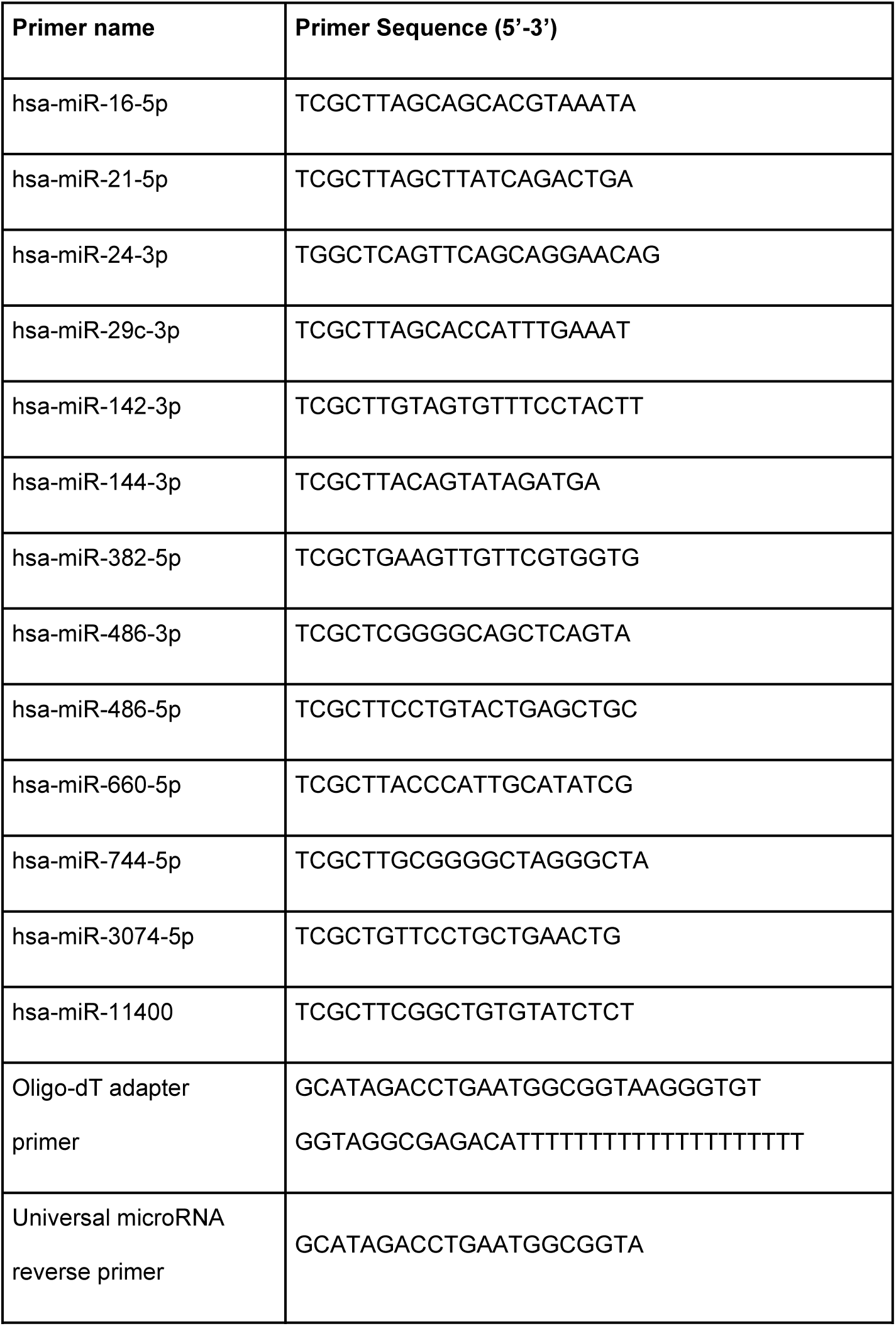
List of primers for microRNA RT-qPCR.

**Supplemental Table 2.** Four microRNAs prioritized in early RA versus early OA and non-OA/RA. For directionality, “Up” reflects upregulation while “Down” reflects downregulation in early RA versus early OA or non-OA/RA. For logFC, >0 reflects upregulation while ≤0 reflects downregulation in early RA versus early OA or non-OA/RA. For FDR, ≤0.05 represents statistically significant differential expression in the pairwise comparison. For proportion, values represent the percentage of samples with logCPM either greater than (for upregulated microRNAs) or less than (for downregulated microRNAs) the median logCPM of the comparison group.

| MicroRNA | Directionality<br>in Early RA | Early RA versus Early OA |  |  |  | Early RA versus Non-OA/RA |  |  |  |
| --- | --- | --- | --- | --- | --- | --- | --- | --- | --- |
|  |  | logFC | logCPM | FDR | Proportion | logFC | logCPM | FDR | Proportion |
| hsa-miR-11400 | Up | 3.27 | 4.81 | 0.02 | 1.00 | 0.03 | 5.95 | 0.99 | 0.83 |
| hsa-miR-21-5p | Up | 1.26 | 10.50 | 0.00 | 1.00 | 0.28 | 10.96 | 0.41 | 0.83 |
| hsa-miR-660-5p | Down | -0.95 | 7.76 | 0.00 | 1.00 | -0.46 | 7.51 | 0.19 | 0.83 |
| hsa-miR-29c-3p | Down | -0.77 | 8.35 | 0.02 | 1.00 | -0.55 | 8.30 | 0.19 | 0.83 |

**Supplemental Table 3.**
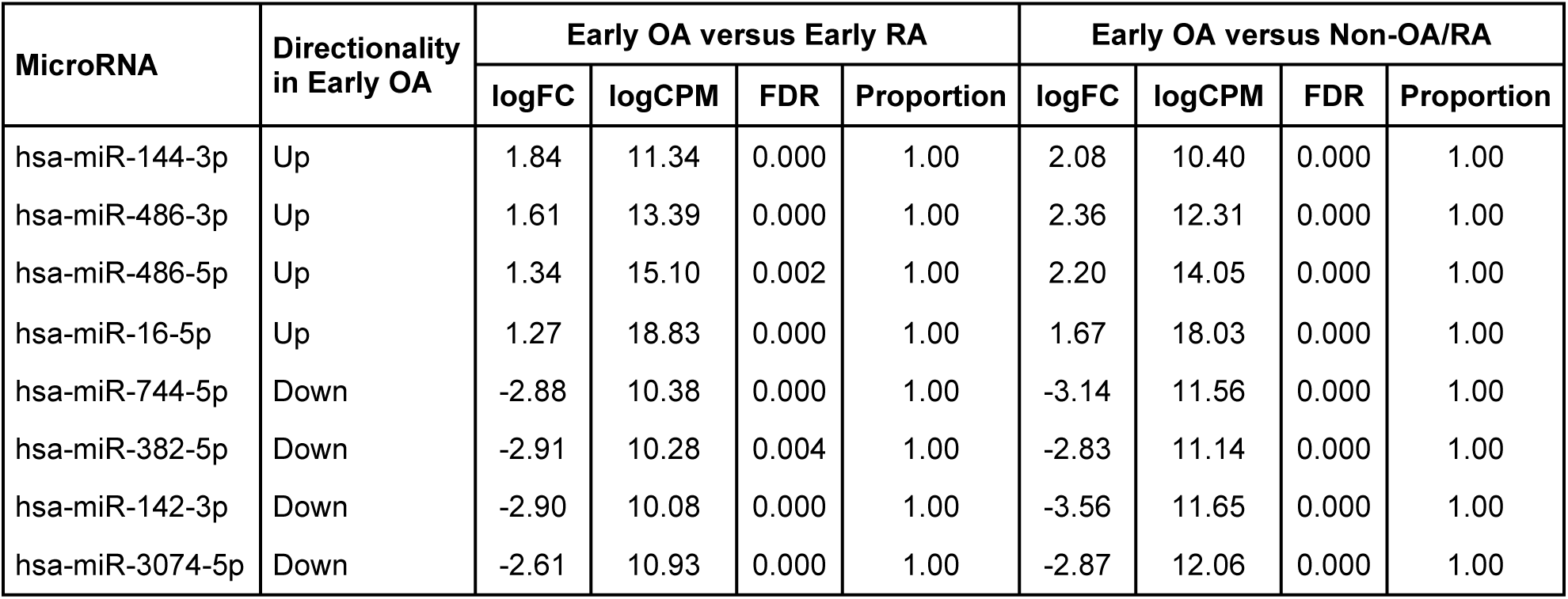
Eight microRNAs prioritized in early OA versus early RA and non-OA/RA. For directionality, “Up” reflects upregulation while “Down” reflects downregulation in early OA versus early RA or non-OA/RA. For logFC, >0 reflects upregulation while ≤0 reflects downregulation in early OA versus early RA or non-OA/RA. For FDR, ≤0.05 represents statistically significant differential expression in the pairwise comparison. For proportion, values represent the percentage of samples with logCPM either greater than (for upregulated microRNAs) or less than (for downregulated microRNAs) the median logCPM of the comparison group.

**Supplemental Table 4.** Characterization of the two study groups used for microRNA RT-qPCR. N = sample size; SD = standard deviation; BMI = body mass index.

| Cohort | Early OA | Early RA | p-value |
| --- | --- | --- | --- |
| N | 8 | 6 | - |
| Sex (female)<br>N (%) | 2<br>(25) | 2<br>(33.3) | 1 |
| Race (white)<br>N (%) | 8<br>(100) | 5<br>(83.3) | 0.89 |
| Age (years)<br>Mean (SD) | 24<br>(5.3) | 63<br>(9.4) | <0.05 |
| BMI (kg/m<sup>2</sup>)<br>Mean (SD) | 30<br>(8.5) | 26<br>(2.9) | 0.30 |

